# Association of Body Anthropometry and Obstructive Sleep Apnea in Children: Impact of Hispanic Ethnicity

**DOI:** 10.1101/2021.10.20.21265294

**Authors:** Emily B Bhattacharjee, Xiaoying Sun, Atul Malhotra, Kelan G Tantisira, Jeremy S Landeo-Gutierrez, Sonia Jain, Rakesh Bhattacharjee

## Abstract

**Objectives:** Obesity is a risk factor for obstructive sleep apnea (OSA) in children. Childhood obesity rates vary amongst different ethnic groups. Here we explore the interaction of Hispanic ethnicity and obesity on OSA risk.

**Methods:** Retrospective cross-sectional analysis of consecutive children undergoing polysomnography and anthropometry using bioelectrical impedance from 2017-2020. Demographics obtained from the medical chart. Children who had also undergone cardiometabolic testing were identified to assess the relationship of cardiometabolic markers with OSA and anthropometry.

**Results:** Data from 1217 children revealed Hispanic children were more likely to have moderate-severe OSA (36.0%) compared to Non-Hispanic children (26.5%), p<0.001. Hispanic children had greater BMI, BMI percentile and percent body fat, p<0.0001. In children that underwent cardiometabolic testing, Hispanic children had significantly greater serum ALT levels. Following adjustment of age and sex, Hispanic ethnicity was not found to moderate the association of anthropometry with OSA, anthropometry with cardiometabolic markers, and OSA with cardiometabolic markers.

**Conclusions:** OSA is more likely in Hispanic children; this relationship is likely driven by obesity status rather than ethnicity. Among children undergoing cardiometabolic testing, Hispanic children were observed to have greater ALT concentrations however ethnicity did not impact the association of anthropometry and ALT or other cardiometabolic markers.

## Introduction

Obstructive Sleep Apnea (OSA) is a common condition in which normal sleep is disrupted by episodic collapse of the upper airway causing sleep fragmentation and abnormal gas exchange, including intermittent oxygen desaturation. In children, the reported estimated prevalence of OSA is about 2-3%(1, 2, 3, 4). While hypertrophy of adenotonsillar tissue comprises as the major risk factor for OSA in children(4, 5), particularly young children, the surge in childhood obesity(6) has resulted in an increase in prevalence with up to 6% of all children being affected by OSA(7, 8, 9, 10, 11). Furthermore, the degree of adenotonsillar hypertrophy necessary for airway obstruction during sleep is significantly less in obese children(12). In context of the high prevalence of childhood obesity, OSA should now be considered a common disease of childhood.

Obesity impacts sleep related breathing via independent mechanisms. Fatty deposits in the anterior neck encroaches the upper airway, narrowing the diameter of the upper airway leading to pharyngeal collapsibility during sleep. In addition, abdominal obesity restricts normal diaphragmatic expansion, particularly during supine sleep, thereby reducing lung volumes and predisposing children to gas exchange abnormalities typical of OSA.

In children, studies have implicated OSA in neurocognitive and behavioral consequences(13, 14, 15, 16, 17, 18, 19) and recently cardiovascular morbidity(20) in children. Of note, similar to childhood obesity, OSA has also been associated with several metabolic derangements in children. Lipid profiles obtained from pre-pubertal children with OSA or adenotonsillar hypertrophy correlate with elevations in low-density lipoprotein cholesterol levels, and reductions in high-density lipoprotein cholesterol levels in children with OSA which reversed following treatment(21). Similar findings have been corroborated in post-pubertal children(22). While it is noteworthy that obesity is a significant risk factor for childhood metabolic disease, the coexistence of OSA in many obese children may augment metabolic disease even further. Adding to the complexity of this relationship, studies have suggested that sleep restriction or lack of sleep(23, 24) and sleep fragmentation induced by OSA(25) may increase obesity risk in children.

Ethnicity is a well ascribed major risk factor for childhood obesity(26), and impacts OSA prevalence(4, 27, 28, 29, 30) in children. Notwithstanding few studies have addressed the influence of Hispanic ethnicity on OSA(31). Here we evaluate a large population of children undergoing both sleep study testing and body anthropometric measurements. We seek to assess the influence of Hispanic ethnicity on the prevalence of OSA as well as body anthropometry, and evaluate the interaction of Hispanic ethnicity with anthropometric measures on OSA. In addition, we evaluated those patients that had undergone cardiometabolic testing within one year of their sleep study to evaluate the relationship of OSA and anthropometry on cardiometabolic maskers, and also assess the influence of Hispanic ethnicity status.

## Methods

All study data were collected retrospectively through approval by the Institutional Review Board at the University of California, San Diego (#180458). Criteria for study inclusion consisted of all children (age<18 years) having a diagnostic polysomnogram as well as anthropometry measured using the InBody 570^®^ Body composition Analyzer (Queensland, Australia). All testing was conducted at the Rady Children’s Hospital Center for Healthy Sleep, San Diego, CA from January 2017 to March 2020. Demographic data including age, sex, ethnicity, medical history were extracted from the electronic medical record (EMR). Children undergoing polysomnography for the purpose of positive airway pressure titration were excluded. During the study period, if children had undergone repeat polysomnography to assess sleep following airway surgery, only presurgical polysomnograms were chosen.

Of all patients included in the study, a subset of patients was also identified using the EMR if they had undergone cardiometabolic blood testing within one year of the date of the time of their sleep lab visit. These patients were separately analyzed to evaluate the relationship of sleep apnea or anthropometry on cardiometabolic profiles.

Overnight laboratory diagnostic PSGs were conducted and scored by sleep technologists and interpreted by a pediatric sleep medicine physician according to American Academy of Sleep Medicine (AASM) criteria(32). Polysomnogram parameters examined included the total sleep apnea hypopnea index (AHI), obstructive apnea-hypopnea index (OAHI), oxygen desaturation index (ODI), and oxygen saturation nadir during sleep. Normal, mild, and moderate/severe OSA was defined as having an OAHI less than 1.5, 1.5 to ≤ 5, and >5 events/hour respectively.

Body mass index (BMI) was calculated using weight and height as measured by a digital stadiometer. BMI percentiles were calculated using CDC normative data in children (ref). Body composition was measured using a bioelectrical impedance analyzer (InBody 570®). The analyzer measures body water through bioelectrical impedance using electrodes that are situated beneath the subject’s feet on the platform and on the palms and thumbs attached to handles on the device. As a result, the device is limited to children who can stand independently. Bioelectrical impedance is used to derive dry lean mass and calculate body fat mass and percentage of body fat. This technique demonstrates strong correlation with dual-energy X-ray absorptiometry scanning(33).

### Statistical analysis

Descriptive statistics were used to summarize demographic characteristics of children. All normally distributed data were presented with sample mean ± standard deviation, data that were not normally distributed were presented with median and interquartile range. Group comparisons between Hispanic and Non-Hispanic children used Wilcoxon rank sum test for continuous variables and Fisher’s exact test for categorical variables. Logistic regression model was used to assess whether the association of each anthropometric variable or cardiometabolic measure with the presence of OSA differed between Hispanic vs Non-Hispanic children. Linear regression model was used to study whether Hispanic ethnicity moderated the association between each anthropometric variable and cardiometabolic outcome. To minimize data skew, cardiometabolic outcomes underwent log transformation in these analyses. All models adjusted for age and sex. Analyses were conducted in R (version 3.6.1).

## Results

Anthropometric and sleep study data were available from 1333 children. Patients who had sleep studies less than 240 minutes were deemed as unsuccessful sleep studies and were thereby excluded from the analysis (n=99). 13 subjects were excluded due to a current age of greater than 18 years. 4 subjects were excluded due absence of ethnicity data. A total of 1217 children (509 females or 41.8%) during the study period were included. Demographic sleep study characteristics are outlined in table 1. Of the study population, 659 (54.1%) children were of Hispanic ethnicity. There were no significant differences in sex distribution between Hispanic and Non-Hispanic children. There was however a statistically significant difference in age with the mean±sd age of Hispanic children found to be 10.9±3.5 years compared to 10.4±3.9 years in Non-Hispanic children (p=0.0043), although this difference is unlikely to be clinically significant.

**Table 1.** Demographic Characteristics and Sleep Apnea outcomes of Study Population

|  | Total<br>(n=1217) | Hispanic<br>(n=659) | Non-Hispanic<br>(n=558) | p Value |
| --- | --- | --- | --- | --- |
| **Sex** |  |  |  | >0.999 |
| Female | 509 (41.8%) | 276 (41.9%) | 233 (41.8%) |  |
| Male | 708 (58.2%) | 383 (58.1%) | 325 (58.2%) |  |
| **Age (years) [Mean±SD, range]** | 10.6±3.7 [1-18] | 10.9±3.5 [1-18] | 10.4±3.9 [3-18] | **0.0043** |
| **AHI (events/hr) [Median, IQR]** | 3.2(1.5-8.3) | 3.8(1.6-10.3) | 2.6(1.3-6.6) | **<0.0001** |
| **OAHl (events/hr) [Median, IQR]** | 2.3(0.9-7.1) | 3.0(1.1-8.7) | 1.9(0.7-5.4) | **<0.0001** |
| **ODI (events/hr) [Median, IQR]** | 0.7(0.2-2.4) | 0.8(0.2-3.0) | 0.6(0.2-1.8) | **<0.0001** |
| **SaO2 Nadir (%) [Median, IQR]** | 93.0 (90.0-94.0) | 92.0(89.0-94.0) | 93.0 (90.0-95.0) | **<0.0001** |
| **OSA Category [Number, %]** |  |  |  | **<0.001** |
| Normal (OAHl<1.5) | 472 (38.8%) | 227 (34.5%) | 245 (43.9%) |  |
| Mild (1.5≤OAHl<5) | 360 (29.6%) | 195 (29.6%) | 165 (29.6%) |  |
| Moderate - Severe (OAHl≥5) | 385 (31.6%) | 237 (36.0%) | 148 (26.5%) |  |

Table Legend – AHI – apnea hypopnea index; OAHl – obstructive apnea hypopnea Index; ODI – oxygen desaturation Index;
SaO2 – oxygen saturation; OSA – obstructive sleep apnea

The median (IQR) AHI and OAHI were 3.2 (1.5-8.3) events/hr and 2.3 (0.9-7.1) events/hr of the entire population. Hispanic children were observed to have significantly larger median AHI and OAHI values at 3.8 events/hr and 3.0 events/hr compared to Non-Hispanic children 2.6 events/hr and 1.9 events/hr (Table 1) (p<0.0001 for both measures. In addition, Hispanic children were observed to have significantly larger median ODI levels and significantly lower median oxygen saturation nadirs. 472 (38.8%) of the 1217 children were observed to have normal sleep studies (OAHI≤1.5 events/hr), 360 children (29.6%) found to have mild OSA (1.5<OAHI≤5 events/hr) and 385 children (31.6%) were found to have moderate-severe OSA (OAHI>5 events/hr). Hispanic children were more likely to have moderate-severe OSA (237 of 659 children or 36.0%) compared to Non-Hispanic children (148 of 558 children or 26.5%), and less likely to have normal sleep studies (227 of 659 children or 34.5%) compared to Non-Hispanic children (245 of 558 children or 43.9%), p<0.001.

Anthropometric measures revealed that Hispanic children were found to have greater BMI, BMI percentiles, body fat mass, and percent body fat (Table 2). However, despite the observed larger polysomnographic indices suggestive of OSA and anthropometric indices in Hispanic children, following adjustment for age and sex, there was no significant interaction of Hispanic ethnicity on the relationship of body anthropometry and the presence of moderate-severe OSA (OAHI>5 events/hr) (Table 3).

**Table 2.** Anthropometric Measures of Study Population

| Anthropometric Measure | Total<br>(n=1217) | Hispanic<br>(n=659) | Non-Hispanic<br>(n=558) | pValue |
| --- | --- | --- | --- | --- |
| **BMI** | 23.7±7.9 | 26.0±8.0 | 21.0±6.7 | **<0.0001** |
| **BMI Percentile** | 66.0±34.7 | 75.9±31.2 | 54.4±35.0 | **<0.0001** |
| **Dry Lean Mass** | 21.4±8.90 | 22.2±8.8 | 20.6±9.1 | **<0.0001** |
| **Body Fat Mass** | 44.5±36.8 | 55.6±38.0 | 31.4±30.5 | **<0.0001** |
| **Percent Body Fat** | 30.9±14.1 | 36.2±12.3 | 24.6±13.3 | **<0.0001** |

Table Legend – BMI – body mass index, Mean ±SD was reported.

**Table 3.** Associations between Anthropometry and OSA (Moderate-severe (OAHI>5) versus Normal (OAHI<1.5)) and Interaction of Hispanic Ethnicity

| Anthropometric Measure | Ethnicity | Odds Ratio (OR) | OR (lower limit) | OR (upper limit) | p Value | Interaction p Value |
| --- | --- | --- | --- | --- | --- | --- |
| **BMI** | Non-Hispanic | 2.055 | 1.559 | 2.708 | <0.001 | 0.709 |
|  | Hispanic | 2.191 | 1.746 | 2.749 | <0.001 |  |
| **BMI Percentile** | Non-Hispanic | 1.478 | 1.195 | 1.827 | <0.001 | 0.876 |
|  | Hispanic | 1.513 | 1.223 | 1.873 | <0.001 |  |
| **Dry Lean Mass** | Non-Hispanic | 1.721 | 1.249 | 2.371 | <0.001 | 0.591 |
|  | Hispanic | 1.868 | 1.392 | 2.506 | <0.001 |  |
| **Body Fat Mass** | Non-Hispanic | 2.019 | 1.529 | 2.666 | <0.001 | 0.811 |
|  | Hispanic | 2.104 | 1.664 | 2.659 | <0.001 |  |
| **Percent Body Fat** | Non-Hispanic | 1.860 | 1.477 | 2.342 | <0.001 | 0.474 |
|  | Hispanic | 2.093 | 1.653 | 2.650 | <0.001 |  |

Table Legend - Separate logistic regression models were performed to assess whether the association of each anthropometric measure (in standardized scale) with OSA (defined as moderate-severe if OAHI>5/hr vs normal if OAHI≤1.5/hr) was moderated by ethnicity status (Hispanic vs non-Hispanic). All models adjusted for age and sex. OR represents the odds of having moderate-severe OSA per standardized unit increase in each anthropometric measure. Interaction p Value indicates whether the association differs between Hispanic and Non-Hispanic children.
*Table Legend – BMI – body mass index*

In the subset of patients who undergone cardiometabolic evaluation within a year of their sleep study, there were no significant differences observed in triglyceride and total cholesterol levels (Table 4) when comparing Hispanic to Non-Hispanic children. There were no differences in inflammation detected as determined by C-reactive protein or the percentage of Hemoglobin A1c. There was however a significant elevation in alanine aminotransferase levels (ALT) in Hispanic children 41.4±37.5 U/L compared to 31.4±21.5 U/L in Non-Hispanic children (p<0.0001).

**Table 4.** Cardiometabolic Measures of Study Patients Having Undergone Blood Testing

|  | Total | n | Hispanic | n | Non-Hispanic | n | p Value |
| --- | --- | --- | --- | --- | --- | --- | --- |
| **Triglycerides (mg/dL)** | 110(85-156) | 205 | 112(88-157) | 136 | 103(78-150) | 69 | 0.153 |
| **Total Cholesterol (mg/dL)** | 155(131-177) | 204 | 151(129-177) | 136 | 158(136-176) | 68 | 0.297 |
| **C-Reactive Protein (mg/L)** | 1(0.5-3.9) | 89 | 1.2(0.5-3.9) | 53 | 0.6(0.5-3.2) | 36 | 0.263 |
| **ALT (U/L)** | 29(21-42) | 365 | 32(23-45) | 220 | 26(19-34) | 145 | **<0.0001** |
| **HbA1c (%)** | 5.4(5.1-5.5) | 106 | 5.4(5.2-5.5) | 74 | 5.4(5.1-5.5) | 32 | 0.531 |

Table Legend – ALT – alanine aminotransferase; HbA1c – hemoglobin A1c, Median (IQR) was reported.

Following adjustment for age and sex, there were no statistically significant differences observed in any of the cardiometabolic markers of focus in those children with normal sleep studies compared to those children with moderate-severe OSA, regardless of Hispanic ethnicity (Table 5). Hispanic ethnicity was also not found to significantly moderate the association between anthropometric measures and any of the cardiometabolic markers (Table 6).

**Table 5.**
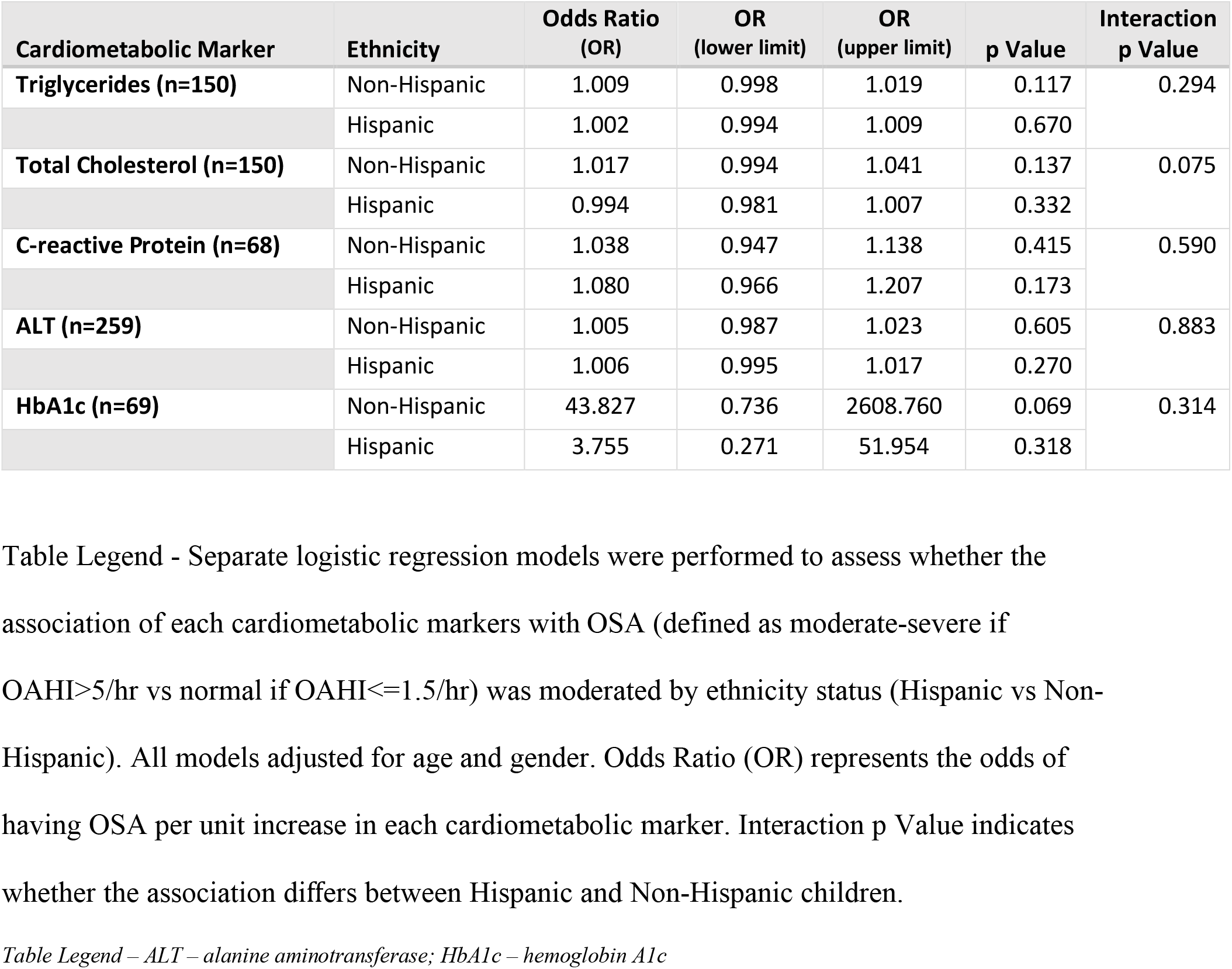
Associations between Cardiometabolic Measures and OSA (Moderate-severe (OAHI>5) versus Normal (OAHI<1.5)) and Interaction of Hispanic Ethnicity

**Table 6.**
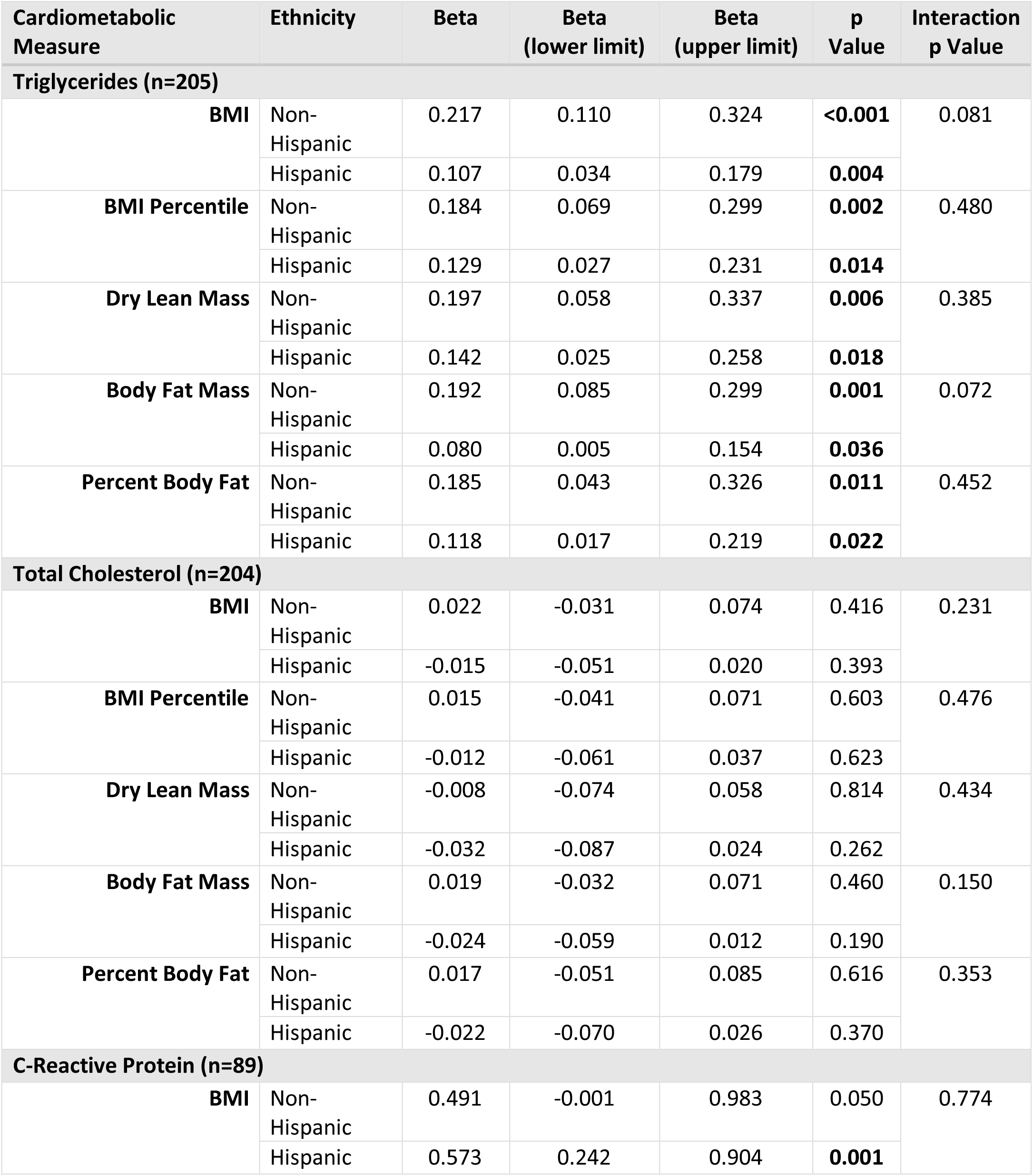
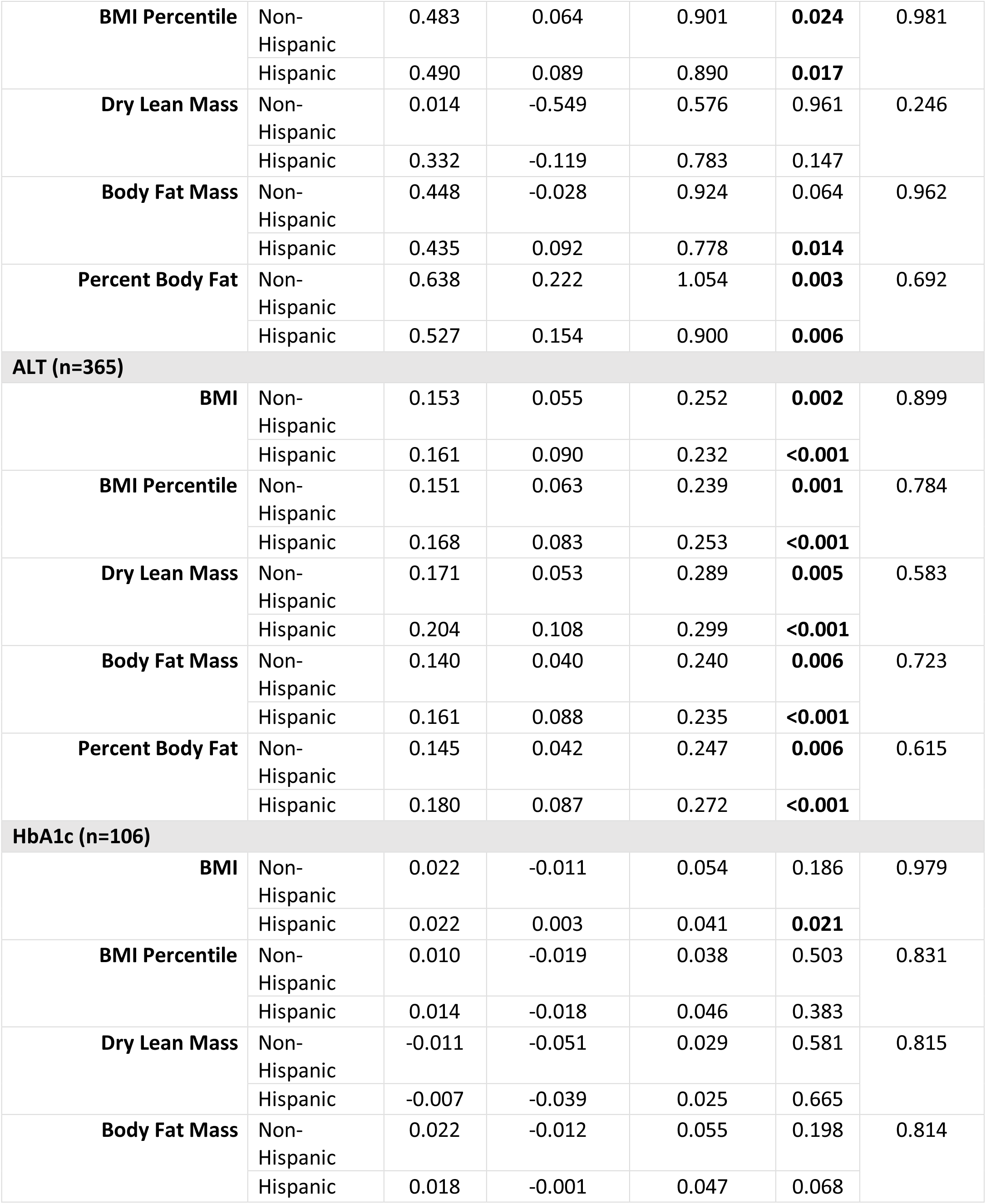
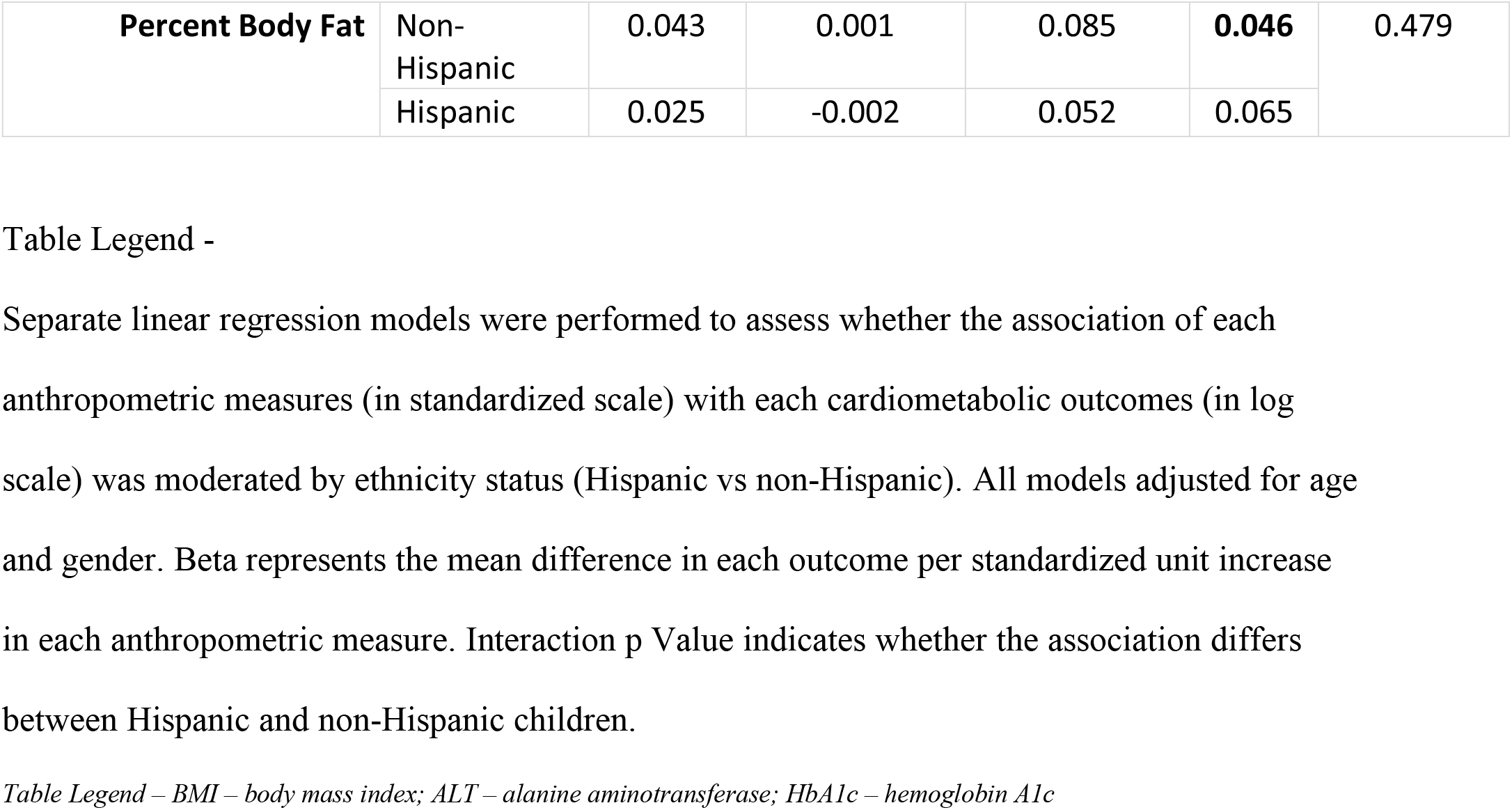
Associations between Anthropometric Measures and Cardiometabolic Markers Interaction of Hispanic Ethnicity

## Discussion

Our findings reveal that in a large clinical population of children undergoing polysomnography, Hispanic children are more likely to have moderate-severe OSA, more likely to have anthropometric measures that suggest obesity, and finally a greater likelihood of having elevated serum ALT levels. However, following adjustment for age and sex, Hispanic ethnicity did not moderate the relationship of OSA and body anthropometry. Notwithstanding, these findings imply that obesity is more common in Hispanic children and given the propensity towards obesity in Hispanic children, they are at greater risk for OSA. We view these findings as important motivation for preventative strategies including diet and exercise in all children particularly in those of Hispanic ethnicity. Moreover, we hope that our findings will improve awareness and recognition of OSA and OSA related complications in Hispanic children.

In addition to the findings on OSA, our study did reveal differential susceptibilities of cardiometabolic risk namely ALT, a surrogate for hepatic steatosis, in Hispanic children. However, following adjustment for age and sex, Hispanic ethnicity did not significantly moderate the association of anthropometry and ALT or other cardiometabolic markers of interest. A recent study of Korean children(34), has also reported elevation of ALT in children with OSA and obesity implying the link of obesity and OSA with cardiometabolic disease, however this study did not assess the impact of different ethnic groups.

Despite our study drawing conclusions from a large dataset of children (n=1217), there are several limitations with our study. First, the relationship of cardiometabolic risk with OSA status and body anthropometry was limited by a smaller sample size, as not all children underwent metabolic blood testing. Further, those children that underwent metabolic blood testing were more likely to have testing done for a specific reason, e.g family history of cardiovascular disease or diabetes risk. Further work will be required to look at the generalizability of our findings. Second, our study did not evaluate hard cardiometabolic outcomes, such as liver biopsy, or cardiac MRI. Thus, we relied on surrogate measures which are typically used clinically. However, we are supportive of further efforts to perform more definitive outcome measures. Third, based on the nature of our study design, we evaluated a clinical sample with an inherent referral bias. Because our goal was to perform a real-world clinical study, we view the findings as important. Nonetheless, we are supportive of efforts to evaluate community cohorts which may provide additional insights. Fourth, we did not evaluate specific phenotypical traits associated with OSA that may be more prevalent in certain ethnic groups such as mandibular cortical width (35) or neck circumference (36). Fourth, our findings are correlative, but we hope they provide supportive data to inform the proper design of rigorous randomized clinical trials to evaluate further the impact of treatment such as positive airway pressure on ethnically diverse children with OSA.

## Conclusions

Data from a large population of children show that prevalence rates of OSA and anthropometric measures of obesity are significantly higher in Hispanic children compared to Non-Hispanic children. Following adjustment, Hispanic ethnicity however did not moderate the association of anthropometry and OSA, OSA and cardiometabolic risk and finally anthropometry and cardiometabolic risk. These findings imply that while the clinical presentation and potential cardiometabolic consequences may vary when comparing Hispanic to Non-Hispanic children, the effect is largely driven by the presence of obesity rather than ethnicity specifically.

## Data Availability

All data produced in the present work are contained in the manuscript

